# Real-time detection of spoken speech from unlabeled ECoG signals: A pilot study with an ALS participant

**DOI:** 10.1101/2024.09.18.24313755

**Authors:** Miguel Angrick, Shiyu Luo, Qinwan Rabbani, Shreya Joshi, Daniel N. Candrea, Griffin W. Milsap, Chad R. Gordon, Kathryn Rosenblatt, Lora Clawson, Nicholas Maragakis, Francesco V. Tenore, Matthew S. Fifer, Nick F. Ramsey, Nathan E. Crone

## Abstract

*Objective*. Brain-Computer Interfaces (BCIs) hold significant promise for restoring communication in individuals with partial or complete loss of the ability to speak due to paralysis from amyotrophic lateral sclerosis (ALS), brainstem stroke, and other neurological disorders. Many of the approaches to speech decoding reported in the BCI literature have required time-aligned target representations to allow successful training – a major challenge when translating such approaches to people who have already lost their voice. *Approach*. In this pilot study, we made a first step toward scenarios in which no ground truth is available. We utilized a graph-based clustering approach to identify temporal segments of speech production from electrocorticographic (ECoG) signals alone. We then used the estimated speech segments to train a voice activity detection (VAD) model using only ECoG signals. We evaluated our approach using held-out open-loop recordings of a single dysarthric clinical trial participant living with ALS, and we compared the resulting performance to previous solutions trained with ground truth acoustic voice recordings. *Main results*. Our approach achieves a median error rate of around 0.5 seconds with respect to the actual spoken speech. Embedded into a real-time BCI, our approach is capable of providing VAD results with a latency of only 10 ms. *Significance*. To the best of our knowledge, our results show for the first time that speech activity can be predicted purely from unlabeled ECoG signals, a crucial step toward individuals who cannot provide this information anymore due to their neurological condition, such as patients with locked-in syndrome. *Clinical Trial Information*. ClinicalTrials.gov, registration number NCT03567213.

## Introduction

Several neurological disorders, including amyotrophic lateral sclerosis (ALS), can result in severe paralysis and loss of speech, having devastating effects on the quality of life of affected individuals. Recent advances in implantable Brain-Computer Interfaces (BCIs) have raised hope for the restoration of communication in this clinical population^1,2^ by utilizing neural activity acquired directly from the cerebral cortex to control a neuroprosthetic device that produces text^3–7^ or synthesizes speech^7–13^. Those BCIs are currently trained using supervised learning paradigms where neural activity is mapped onto target representations^14,15^, such as phonemes or acoustic units, and are therefore dependent on accurate temporal alignments to achieve proper outputs. For this reason, many prior studies in the field have relied on datasets collected from patients who had normal speaking capabilities, such as epilepsy patients^8,13,16,17^ or patients who underwent surgery for glioma removal^9,11^ – datasets where the temporal alignment can be obtained from simultaneous acoustic recordings.

In recent years, clinical trials have begun exploring the extent to which approaches previously used in normal speaking subjects can be translated to people in actual need for such a technology^3,7,18,19^, and while those enrolled clinical-trial participants were speech impaired, their diseases had not yet been progressed into a state of total paralysis that prevented inferring such an alignment. However, in cases where the disease has already progressed to the locked-in syndrome (LIS)^20,21^, it may not be possible to infer the temporal alignment at all from acoustic data. In pioneering work by Guenther et al.^22^, a participant living with LIS was able to accurately synthesize vowels continuously using a Kalman filter-based decoding approach with closed-loop neurofeedback. Additionally, more recent work by Chaudhary et al. gave a completely locked-in patient a novel means of communications by spelling sentences using a paradigm that required modulating firing rates with respect to auditory feedback^23^.

In this study, we make a first step toward acoustic-free model training by assuming that no temporal alignment can be obtained from simultaneous microphone recordings. For this early work, we focus only on localizing and identifying neural activity related to speech processes. Voice Activity Detection (VAD) systems play a crucial role in acoustic speech processing fields, such as automatic speech recognition^24^ or speaker diarization^25^, where they may be used in early processing stages to exclude non-speech data when computing acoustic features or embedding vectors. Similarly, many recent BCI studies have also utilized approaches to locate and isolate neural activity related to speech production in their pipelines as an intermediary step to constrain the solution space of speech decoding tasks, both for word recognition^19,26^ and synthesis applications^18^. Another application for these neural Voice Activity Detection (nVADs) systems of particular relevance to BCIs is to prevent leakage of speech-related activity into computation of baseline statistics within real-time systems. Decoding performance can degrade over time because the feature space may shift linearly beyond the range expected from the training data. nVAD techniques could help here to determine which parts of the neural data should be considered when updating a running baseline, rather than relying only on a fixed time window containing both speech and non-speech activity. To the best of our knowledge, all previous methods have relied on supervised learning machines trained directly on acoustic ground truth^18,19,27^ or labeled information^26^ inferred from behavioral cues^28^ – suggesting that such approaches may not translate to individuals where their disease does not allow vocalization or any observable articulatory movements.

Here, we present first results on unsupervised detection of neural voice activity from unlabeled ECoG signals. We did so by setting up an experiment in which a clinical trial participant was instructed to read single words, and where the majority of time for each recording session did not carry speech activity – a design decision we actively exploited to automatically assign identified segments as either speech or non-speech classes. We utilized a graph-based clustering approach^29^ to find structural patterns with a fixed temporal context in high-gamma activity extracted from ECoG recordings, and used those estimated clustering labels to train a recurrent neural network (RNN). In our evaluation, we first quantified the alignment error between estimated labels from the clustering approach and ground truth acoustic speech information to determine ranges of expected error rates. Next, we compared the performance of our RNN architecture trained on those estimated labels with respect to baseline models previously proposed in the literature trained on VAD labels inferred from acoustic speech. From here, we then inspected how well our model translated to unseen words.

## Material and Methods

### Participant and experiment design

We conducted an experiment with a clinical trial participant (CC01, male) in his 60s with dysarthria due to ALS, who had been implanted with two ECoG arrays with 64 electrodes each (4-mm center-to-center spacing, 2-mm diameter) covering speech and upper-limb cortical areas (Figure 1**a**). The participant could speak, but his speaking capabilities were limited, and continuous speech was mostly unintelligible due to his neurological condition^18,19^ (speech was rated with 1 point out of 5 on the ALSFRS-R measure^30^). In a speech production task, we presented single words on a monitor in front of him and gave instructions to read them out loud. For each trial, the target word was presented for 2 seconds following an inter-trial interval of 3 seconds. Overall, the word pool consisted of 50 words^3^, and each word was repeated twice in each session. We repeated this experiment across 10 days over a period of 9 weeks. Furthermore, we also collected single word data from a larger word pool of 688 words, which we used to quantify generalization towards unseen words. In this corpus, each word only appeared once, and none appeared in the training data. At the start of each recording day, we conducted a syllable repetition task, which was used for normalizing the neural data. The syllable repetition task was constant across all days to achieve similar statistics for the baseline, in accordance with a prior publication with the same study participant^18^.

**Figure 1.**
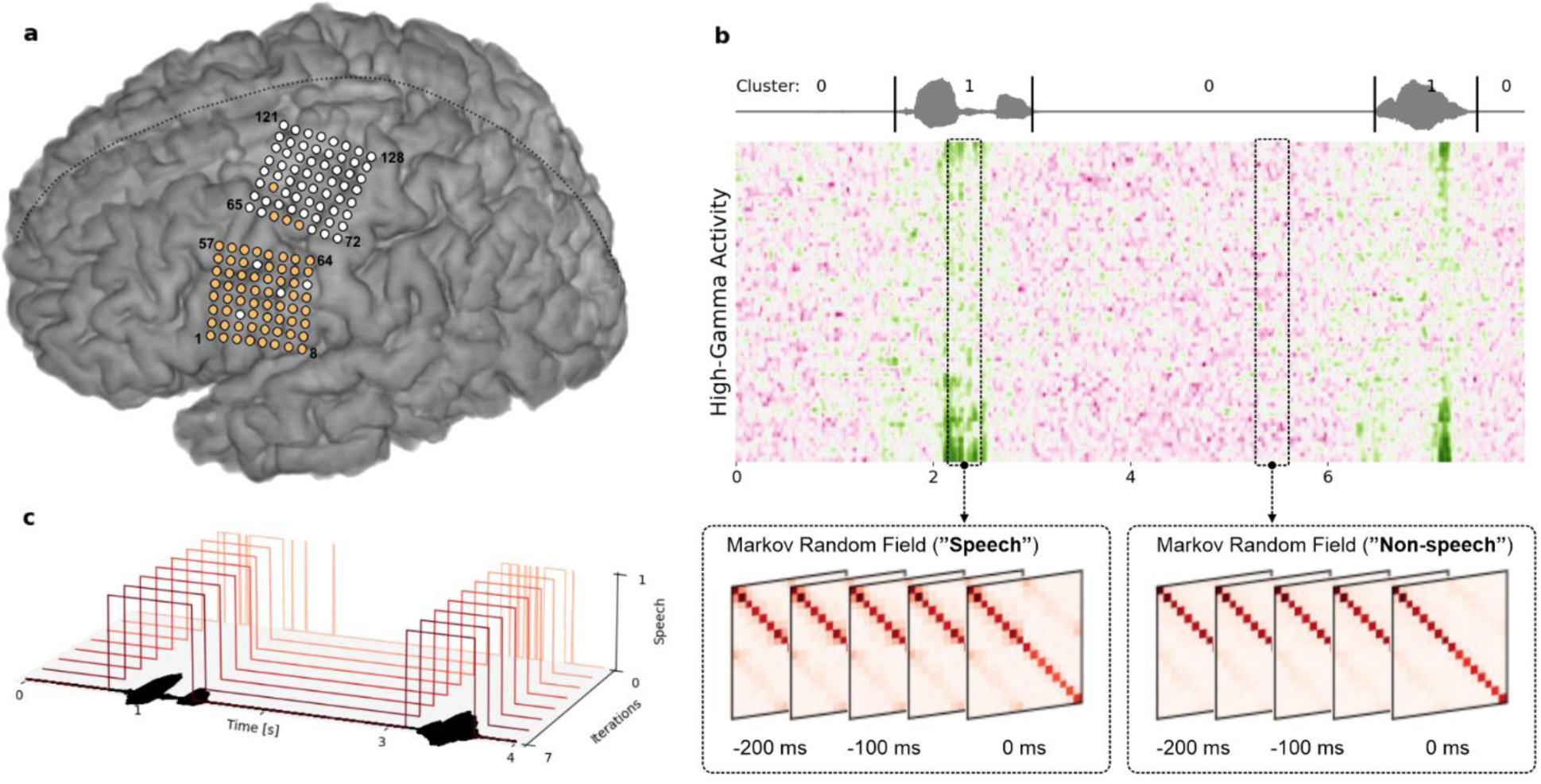
| Overview of the experiment setup and clustering approach. **a** Placement of implanted electrode grids covering speech and upper limb cortical regions. Electrodes highlighted in orange were selected for this study based on previous reported results^18^. **b** Illustration of the TICC clustering approach to identify speech and non-speech segments in each trial using one Markov Random Field per cluster. **c** Visualization of the iterative clustering process of the TICC algorithm, starting from an initial alignment derived from Gaussian mixture clustering, until convergence. The acoustic waveform on the x-axis serves as a reference to the found speech clusters.

Neural data was digitized using a NeuroPort System (Blackrock Neurotech, Salt Lake City, UT, USA) with a sampling rate of 1 kHz. Audio data was recorded at 48 kHz using an external microphone (BETA® 58A, SHURE, Niles, IL). We used BCI2000^31^ for stimulus presentation and for aligning neural and acoustic signals for offline analysis. The clinical trial (ClinicalTrials.gov, NCT03567213) was approved by the Johns Hopkins University Institutional Review Board (IRB) and by the FDA (under an investigational device exemption) to test the safety and preliminary efficacy of a brain-computer interface composed of subdural electrodes and a percutaneous connection to external EEG amplifiers and computers. The participant gave informed consent after being counseled about the nature of the research and implant-related risks, and was implanted with the study device in July 2022.

### Cortical mapping

The positioning of both subdural ECoG grids was determined via anatomical landmarks from pre-operative structural (MRI) and functional imaging (fMRI). After the surgical implantation of the grids, we conducted a post-operative CT scan, which was co-registered to a pre-operative MRI for verification of the anatomical locations of the two grids. Figure 1**a** shows a rendering of the participant’s brain and the locations of both electrode grids, where the 64 electrodes highlighted in orange were relevant in this study with respect to prior observations^18^ about encoded speech activity.

### Signal processing and feature extraction

We obtained speech-related features from raw ECoG signals by extracting the high-gamma (HG) band, which has shown to track closely the location and timing of speech production neural activation^32,33^ and has been successfully employed in previous studies for speech BCIs^4,18,19,34–36^.

First, we removed all bad channels (19, 38, 48 and 52) based on visual inspection and applied a common average referencing (CAR) filter across each grid independently. Next, we selected the top 64 channels with the strongest activation during overt speech production, identified in a previous study^18^ with the same clinical trial participant. We then used a bandpass filter (IIR Butterworth, 4th order) to extract the broadband HG band in the range of 70 – 170 Hz and a notch filter (IIR Butterworth, 4th order) to attenuate the first harmonic of the line noise in the range of 118 – 122 Hz. Finally, for each channel we computed logarithmic power features with respect to a window size of 50 ms and a frame shift of 10 ms. We normalized all features to zero mean unit variance (z-score normalization) with respect to a syllable repetition task conducted at the beginning of each recording day to calibrate the system for day-specific high-gamma changes (see supplementary Figure S1 about the stability of the ECoG signals during the study period). Before using these features for baseline model training, we augmented each frame with a context stacking of 6 consecutive intervals to model temporal dependencies of up to 300 ms in the past. This step was not included in the clustering procedure as the clustering algorithm itself manages a fixed window of past frames to account for the temporal relationships in each cluster.

The acoustic data for performance evaluation was collected at 48 kHz, resampled to 16 kHz and segmented into corresponding windows of 50 ms and 10 ms frameshift to match the alignment with the HG features. We verified that no channels had been contaminated with acoustic artifacts by using Roussel’s method^37^. The details of the contamination report are given in supplementary Figure S2.

### Unsupervised temporal localization of speech production

To identify speech-associated activity in neural recordings, we adopted a graph-based clustering approach named Toeplitz Inverse Covariance-based Clustering (TICC), specifically designed for discovering common subsequences in multivariate time series data. This unsupervised algorithm defines one Markov Random Field (MRF) per cluster and describes relationships in the form of connections between input features. In our study, these connections would describe dependencies between the neural activity of different electrodes, both with respect to spatial and (potentially) causal temporal patterns.

TICC’s training procedure is based on an iterative optimization method that employs a variation of the expectation maximization algorithm which first alternates cluster assignments before updating its cluster parameters. Here, the cluster assignment step is based on the path with the minimum cost, obtained using a dynamic programming paradigm. Once this path has been found, the maximization step updates the cluster parameters based on the assigned data points. The training procedure converges when no data points are assigned to a different cluster and are therefore stationary.

Besides the number of clusters, the TICC algorithm can be configured with respect to the length of the temporal context and regularization parameters. By specifying multiple layers for the MRFs, data points won’t get clustered in isolation but in context to neighboring past observations, allowing it to learn cross-time relationships. Note that temporal layers in the MRFs also obey the Toeplitz constraint to be time-invariant. The regularization parameters β and λ signify the penalty factor for adjacent subsequences being assigned to the same cluster and denote the sparsity level in the MRF’s graph structure characterizing each cluster, respectively. A higher β value will result in a greater likelihood of adjacent subsequences being assigned to the same cluster.

Figure 1b shows an illustration of the TICC clustering approach. Two MRFs segment the high-gamma activity into speech and non-speech classes. In this example, both MRFs have multiple layers to not only draw insights from spatial characteristics but also capture temporal dynamics of up to 200 ms into the past. The gray waveform at the top has been time-aligned to the neural recordings for visual attribution of the high-gamma activity. Although the clustering assignments do not reveal which clusters belong to speech activity due to their unsupervised nature, we can infer cluster classes based on the length of their subsequences – exploiting the setup of the experiment design. Fig 1c visualizes the clustering process for one recording session. The x-axis represents time and shows a snippet of two trials and the acoustic speech signal as a reference guide. The z-axis shows each iteration from the TICC algorithm until convergence, where found cluster alignments are plotted as curves. The y-axis indicates found speech activity. We based our initial alignments (iteration 0) on clusters found by a Gaussian mixture model, and iteratively optimized those using the TICC algorithm.

### Neural voice activity detection approach

We based our nVAD model on the same recurrent neural network architecture from our previous study on synthesizing speech online^18^, originally inspired by the work from Zen et al.^38^ For this binary classification task, all recurrent layers utilize long-short term memory cells^39^ to learn the temporal dynamics across the individual channels. In total, the network architecture comprises three layers: two LSTM layers with 128 units each and one linear layer with 2 output units, resulting in 231,682 internal weight parameters. We used the cross-entropy loss in conjunction with the softmax activation function to estimate the error between network predictions and target labels during network training, and employed Adam^40^ as our optimizer with a learning rate of 0.0001 and trained the architecture for 20 epochs in each fold, while storing the best performing weights in accordance to the minimum validation loss. Network training uses the truncated backpropagation through time (BPTT) algorithm^41^ with hyperparameters *k_1_* and *k_2_* set to 50 frames of high-gamma activity, respectively, such that the unfolding procedure was limited to 50 frames (0.5 s) and repeated every 50 frames (0.5 ms).

### Closed-loop system design

We built a real-time BCI that communicates directly with BCI2000 about any segments identified as speech. This system was implemented on top of *ezmsg*^42^ – a Python framework that facilitates the development of closed-loop streaming applications by enforcing a software architecture composed of a directed acyclic graph structure. Each node in this graph is responsible for a particular self-contained task, such as computing high-gamma features from raw ECoG signals. We used a network of such nodes to perform tasks that receive ECoG signals, compute features, predict voice activity and communicate results back to BCI2000, including logging functionality between all mentioned nodes for evaluation. In the backend, *ezmsg* utilizes asynchronous coroutines to enable concurrent executions of those tasks. Our closed-loop processing pipeline was capable of producing low-latency feedback as the accumulated computational cost did not exceed the frameshift of 10 ms. Communication with BCI2000 was based on ZeroMQ (ZMQ) as a networking abstraction layer.

## Results

### Identification of speech segments

In this study, we only distinguished between speech and non-speech segments in the neural data, so that all words were summarized in one speech cluster. Another potential approach would be to cluster for each stimulus individually, assuming they were known upfront from the experiment design. However, preliminary analyses suggested that resulting clusters per word find less reliable cluster parameters, potentially converging towards clusters that only identify part of speech segments. We hypothesize that this is related to the inherently smaller amount of data and less variability in the neural activity. When clustering for all words, it is not required to know a specific stimulus or the number of stimuli in advance and is thus also suitable for experiment designs where open questions are asked. We obtained ground truth voice activity information from time-aligned acoustic spectrograms of the microphone recordings which were only used to quantify the accuracy of identified speech segments. We based our evaluation metric on the Levenshtein distance to determine the minimum distance between estimated VAD labels and acoustic VAD ground truth, where all operations for changes were assigned a fixed cost of 10 ms.

To infer suitable hyperparameters for the TICC algorithm we utilized ECoG recordings from a single patient with drug-resistant epilepsy (male, between 16-20 years old) who had undergone video-EEG monitoring to localize his seizure onset zone. We particularly chose this data as the implanted ECoG grid covered cortical speech areas similar to our clinical trial participant (see supplementary Figure S3 for details about the grid placement in the epilepsy surgery patient). Note that the electrode grid in the epilepsy surgery patient was implanted in the right hemisphere, yet we were able to measure strong speech-related high gamma activity during speech production. Similar observations have previously been reported in the literature^13^. We ran a grid search across predefined ranges for the β and λ hyperparameters and selected those which achieved lowest alignment errors with respect to ground truth voice activity of the epilepsy patient, leading to a hyperparameter configuration of β = 50 and λ = 11e^-4^.

Our results are summarized in Figure 2a. On a held-out day used to report intermediate results from the clustering algorithm (from now on referred to as development set), we achieved a median alignment error of 530 ms per trial, while 75% of the trials were below 752 ms (average speech duration: 1.2 s). In 8 out of 400 trials, speech could not be detected through the clustering approach and, additionally, 10 trials resulted in alignment errors above the average speech duration of 1.2 s. Figure 2b shows an excerpt of the first 5 trials of the first day in the training set for visual inspection. The top panel visualizes high-gamma activity and how frames have been clustered after applying the TICC algorithm with the same configuration of hyperparameters obtained from the epilepsy patients data. The bottom panel shows the time-aligned speech spectrogram and ground truth VAD information based on the acoustic signal. Overall, the clustering approach can identify consecutive segments of spoken speech reliably in the majority of the cases, leading to labels that can be utilized to train a supervised model that predicts speech activity for an incoming stream of high-gamma frames without calculating the minimum alignment path using dynamic programming strategies.

**Figure 2.**
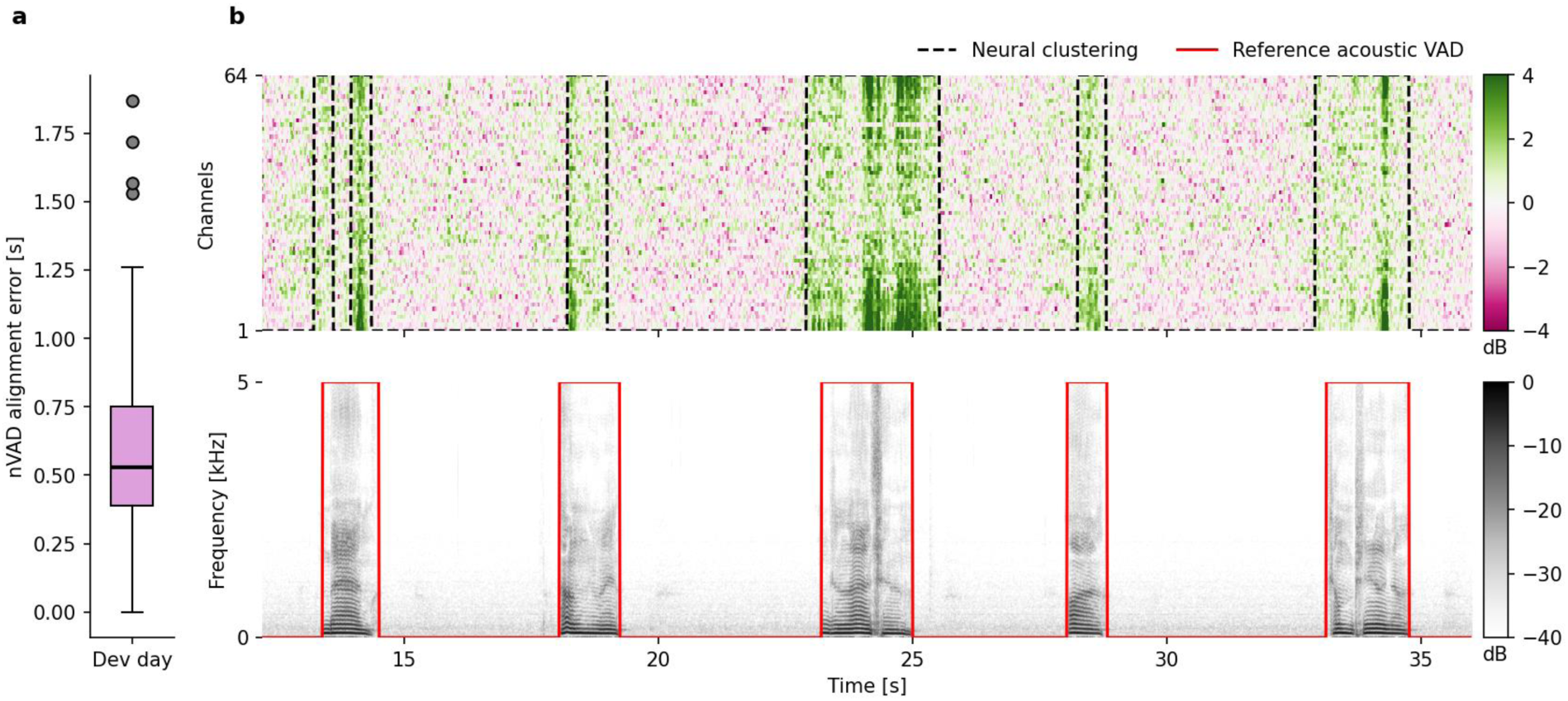
| Comparison between VAD labels estimated from acoustic and high-gamma representations. **a** Minimum alignment error computed via Levenshtein distance between neural speech clusters and acoustic reference VAD using the development set (n = 400 trials). Box indicates boundaries between quartiles Q1 and Q3, and whiskers represent range of data within 1.5 times the interquartile range. Outliers correspond to trials for which no speech clusters could be found from the neural activity. **b** Visual example of the first 5 trials from the first day in the training set. Top panel shows estimated speech clusters using the TICC algorithm (dashed black line) on high-gamma features and bottom panel the corresponding time-aligned speech spectrogram from the acoustics with reference VAD (solid red line).

### Temporal context provides less accurate speech clusters

Next, we analyzed if nVAD labels can be more accurately determined by including causal temporal contextual information. Here, we adapted the TICC algorithm to avoid repetitive information from the 40 ms overlaps in the feature extraction pipeline by adding a dilation hyperparameter indicating the spacing between consecutive high-gamma frames. In accordance with Soroush et al.^27^, we investigated temporal dynamics up to 300 ms into the past. MRFs with only one layer correspond to no context information, with five layers up to 200 ms into the past (as represented in Figure 1b), and with 7 layers of up to 300 ms, where each additional layer introduces a dilation of 5 frames to avoid repetitive information from the 40 ms overlap in the feature extraction pipeline.

Similar to Figure 2a, we report our observations on the development set and used the minimum distance between estimated VAD labels and ground truth labels calculated on the speech spectrogram as the error metric, again with a cost of 10 ms per off-diagonal step in the alignment matrix. Figure 3 visualizes our results in the form of boxplots. We found that the median alignment error increased as more temporal context information was captured in each feature vector. We hypothesize that this trend stemmed from the growing number of features enabling more complex relationships in the spatio-temporal connections, which were inadequately supported by the limited amount of data, leading to increasingly inaccurate cluster parameters. We observed similar results with respect to the data used to determine appropriate hyperparameter choices; therefore, all further analyses were conducted with only one-layer MRFs.

**Figure 3.**
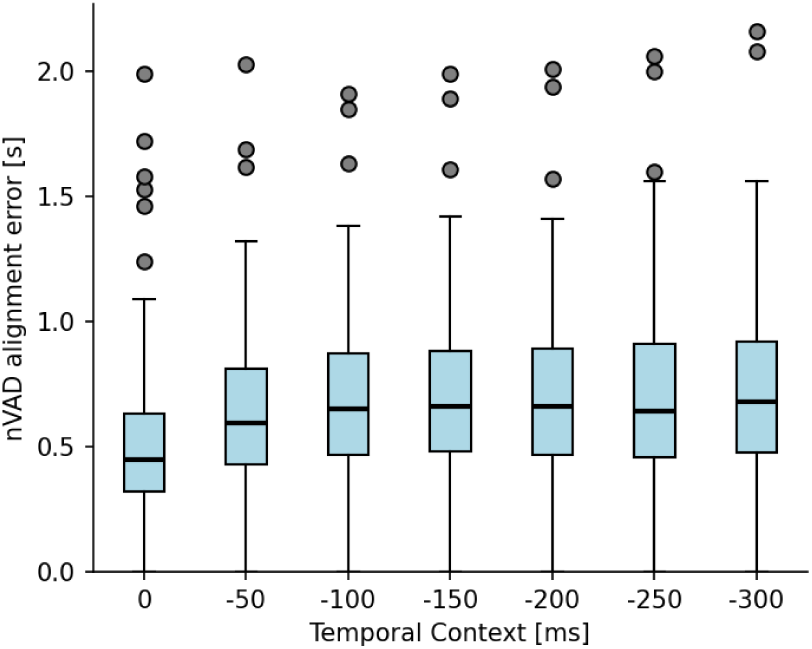
| Adding temporal context leads to less accurate nVAD labels. Trend of inaccuracies for estimated nVAD labels increases with including context information. No context information and 300 ms into the past correspond to MRF’s with number of layers of 1 and 7, respectively. Evaluation was conducted on the development set to report the impact of including temporal context, however, we based our decision on using only one layer in the MRFs on the lowest error score obtained by the TICC algorithm with respect to the data from the epilepsy surgery patient.

### Cluster parameters suggest consistent task-specific activity in motor cortices

The graphical dependency structures underlying cluster representations allow learned relationships to be interpreted and pinpointed to cortical areas known to elicit activity during speech – enabling us to verify that proper representations have been learned. We analyzed the differences between both speech and non-speech MRFs to reveal which connections between electrodes contribute to what extent to the decision-making process. Our findings are visualized in Figure 4. Each circle on the brain plot belongs to one channel. The color of the circle represents how much a particular channel contributed in the decision-making process of the clustering assignment and the size indicates the total sum of the interdependencies between channels. The plot reveals that the differences in high-gamma activity features from electrode channels located in vM1 and dM1 were predominantly used to discriminate between speech and non-speech clusters in the TICC algorithm. Both of these cortical areas have already shown speech activity to various degrees in our prior publication^18^. Moreover, the plot suggest that the algorithm focused on a rather smaller subset of electrodes compared to our prior publication on synthesizing keywords where the supervised nVAD model based its decision on a much broader network of electrodes across motor, premotor and somatosensory cortices. We hypothesize that this is related to the different machine learning approaches (a recurrent neural network compared to the TICC algorithm), the increased number of word stimuli (50 stimuli instead of 6) and the variability in the data as some words in the 50-word corpus are longer and more effortful to articulate.

**Figure 4.**
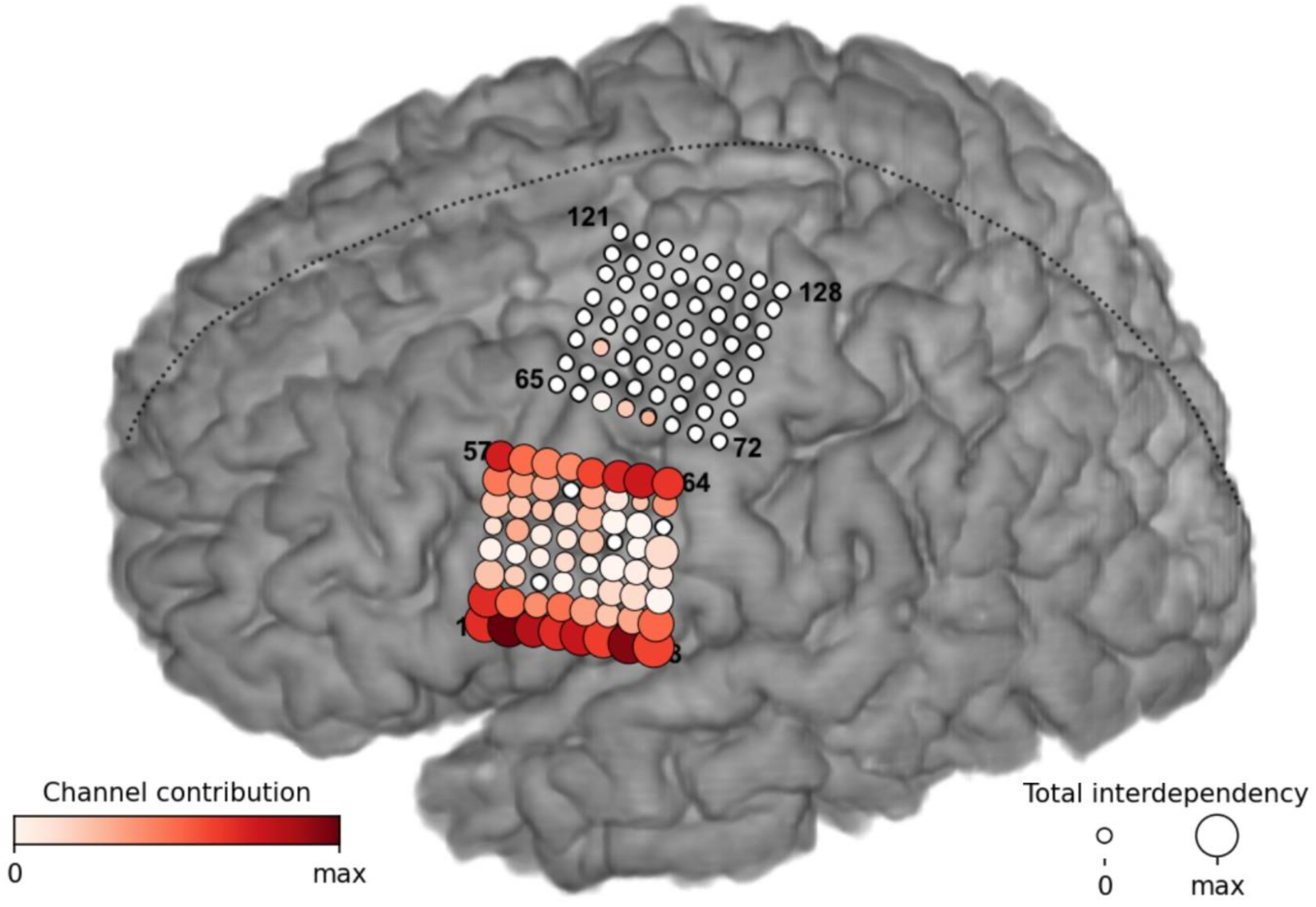
| Cluster assignments mainly driven by differences in inter-electrode connections in vM1 and dM1. Visualization of the differences in the found MRF structures between both speech and non-speech clusters. The color coding of the circles represents electrode contributions, while the size indicates the strength of inter-channel dependencies. These relationships show that the TICC algorithm focused primarily on spatial high-gamma activity patterns between electrodes in vM1 and dM1 when deciding which cluster to assign.

### Predicting speech from neural activity

We evaluated our proposed approach using a leave-one-day-out cross-validation method to quantify model performance across multiple days. Moreover, this prevented day-specific information from the testing days leaking into the training set which may wrongfully bias generalization. We compared our approach trained on the estimated labels from the TICC algorithm against two other methods previously reported in the literature^27^, namely logistic regression (LR) and a LeNet-style convolutional neural network (CNN)^43^, both trained on ground truth information acquired from the acoustic speech spectrogram. For these baseline models, we followed the corresponding study by Soroush et. al.^27^ and used context stacking up to 300 ms into the past, with no additional sequence modeling techniques that would consider outputs from previous time steps.

Our results from the cross-validation are summarized in Figure 5. For each day, we report the alignment errors for our approach (light blue) and the two baseline models (pink and yellow) as boxplots (samples per day: n_1_ = 306, n_2_ = 204, n_3_ = 204, n_4_ = 204, n_5_ = 204, n_6_ = 204, n_7_ = 204, n_8_ = 102, and n_9_ = 204). The dashed red line indicates the average speaking duration of 1.2 s per prompted word from our participant. Across all days from the study period, we observed median error scores between 440 and 645 ms, where 50% of the trial-based errors were in the range of 380 to 710 ms. Furthermore, we also observed that in 5.2% of the trials (96 out of 1836 trials) our model was not capable of detecting speech instances at all or made prediction errors that exceeded the average speaking duration. We excluded those outliers in Figure 5.

**Figure 5.**
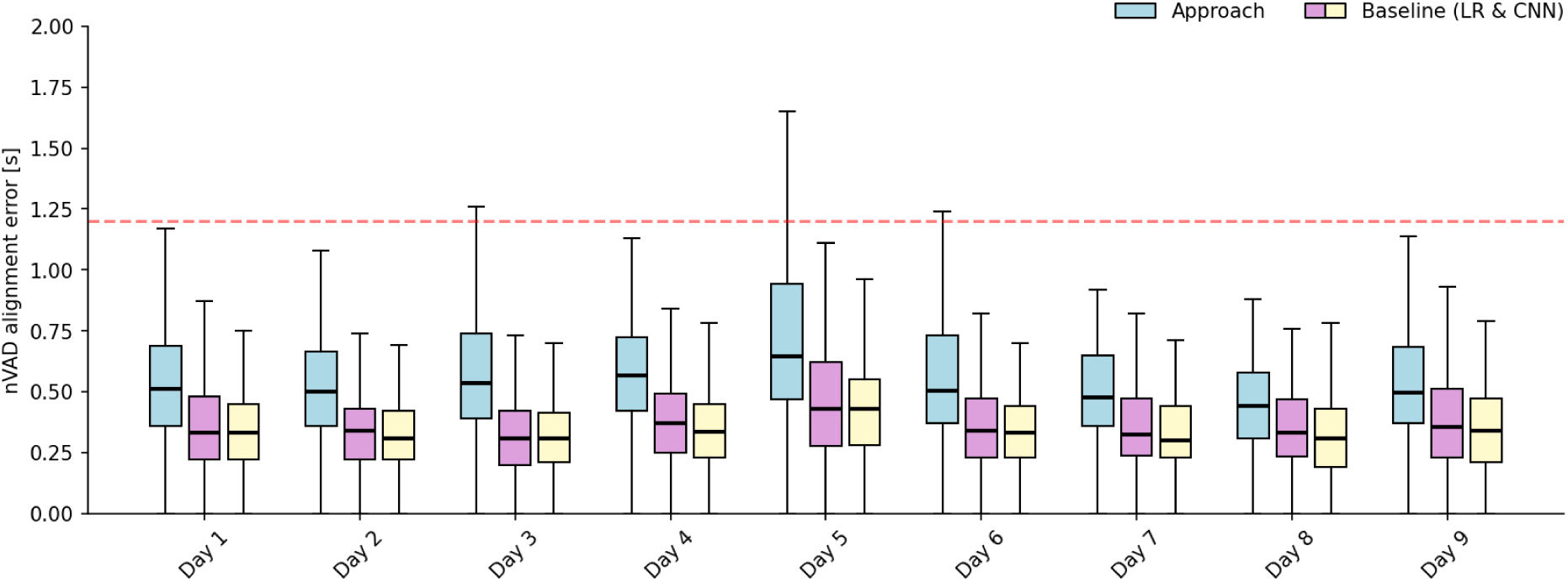
| Cross validation results regarding the proposed approach and baseline models. Alignment errors are reported with respect to the specific held-out day in each fold. Box plots indicate that our approach achieves consistently higher error rates in the range of 140 and 215 ms than models trained on ground truth VAD information.

Regarding the baseline methods, the CNN model achieved overall the lowest alignment errors with median scores between 300 and 430 ms across days, where 50% of the trials deviated between 220 and 450 ms from the ground truth acoustics. For the logistic regression, the alignment errors were slightly higher between 310 and 430 ms in median, which was in line with previous findings on sEEG data^27^. Here, 50% of the trials had alignment errors between 230 and 490 ms.

Although the results from our approach were not on par with baseline models trained on ground truth VAD information, we observed that our approach was still capable of detecting the majority of spoken speech, up to 77% per day, and on average 70% across days. This would be particularly useful for filtering out speech frames during online computations to obtain normalization statistics based on streaming neural activity.

### Generalizability towards unseen words

Next, we analyzed the applicability to spoken words beyond our training corpus of 50-word stimuli to quantify generalization. We recorded an additional corpus of 688 words (each word was only repeated once) across 7 sessions on one particular day (outside of the training days) and computed the mean alignment errors for all trials. The average speaking duration regarding of unseen words was 1.3 s per word. Our results do not show any substantial deviations from those word stimuli that were present in the training corpus. The median alignment errors were between 446 and 490 ms, with 50% of the trials occurring in the 340 and 650 ms range, suggesting that this approach is also applicable to unseen word stimuli.

## Discussion

Here, we demonstrate a BCI that is capable of identifying speech activity in real-time from ECoG signals recorded from speech-related cortical areas in a clinical trial participant living with ALS. Prior studies reporting on voice activity detection from neural activity have relied on ground truth acoustic speech information to train predictive models – a major challenge when translating such findings to paralyzed individuals who have lost their ability to speak. Our approach utilizes a graph-based clustering technique to localize consecutive segments in the neural data related to speech production. We designed an experiment paradigm that can infer which clusters most likely belong to speech activity based on their clustering lengths. By training a recurrent neural network on these estimated alignments, we were able to identify the majority of speech activity in more than 92% of the trials.

While the performance of our approach was not on par with baseline models trained on ground truth acoustic speech information, it would not be reasonable to expect equivalent or better performance in the absence of ground truth. The timing and magnitude of muscular contractions preparing for and executing phonation and articulation do not have a one-to-one correspondence with the timing and magnitude of the acoustic waveform produced by speech, which serves as the ground truth for VAD. Consequently, the timing and magnitude of neural activity in sensorimotor cortex, which form the basis for nVAD, are not expected to be perfectly aligned with spoken acoustics. Moreover, while the signal-to-noise ratio of ECoG high-gamma power modulation has proven sufficient for decoding speech, it is nevertheless non-stationary and dependent on imperfect estimates of its noise floor during non-speech segments, derived here from a separate session with cued speech segments. In spite of these challenges for nVAD, we found that our approach could detect the majority of speech. Analyses on seen and unseen word stimuli revealed that recall scores of approximately 78% could be achieved, compared to 89% from the CNN baseline models. While our current approach was not capable of always isolating each spoken word in its own unique segment, additional postprocessing strategies may help prevent such behavior. Such strategies have been used in the past to correct misclassified frames based on a fixed window of predictions^44^.

By interpreting and comparing cluster parameters, we found that assignments were mainly driven by differences of neural activity in a subset of the electrodes in the vM1 and dM1 cortical regions and their interconnections. Even though many more electrodes show high-gamma activations during overt speech production, the clustering approach converged to similar weights and interconnection weights for both speech and non-speech MRFs on those electrodes. One explanation of this behavior might lie in the high-gamma activity variability across word stimuli, and that the TICC algorithm identified those less reliably when making the binary assignment.

A limitation of our study is that our participant was still able to speak, albeit with significant dysarthria and poor intelligibility. Thus, it remains to be seen if our approach translates to patients who are incapable of producing audible speech. In this study, we focused intentionally on a patient who could still speak so that we could compare the performance of our approach with ground truth speech acoustics and to estimate the extent of alignment errors – which would not have been possible if speech had been absent.

In this pilot study, we addressed the open challenge of training a BCI that identifies speech without having time-aligned neural and acoustic data. Our results show that a graph-based clustering approach can identify segments of spoken speech in neural recordings with median alignment mismatches below 500 ms. Despite this inaccuracy, we were able to train VAD models and deploy them in a real-time streaming scenario to predict speech activity online. The error rate may be small enough for practical application. We believe this would be particularly useful for avoiding the inclusion of speech frames when calculating baseline neural activity during non-speech segments and for real-time gating of speech decoders in speech BCIs, including brain-to-text and brain-to-speech applications. Moreover, our approach could also benefit BCI systems by acting as a switch to toggle on the decoder when the user generates silent speech, and toggle off after some time of silence. This would prevent undesired random speech decoding when the user is doing other tasks that somehow affect motor activity. Future work is necessary to determine whether our approach is equally effective for individuals who can no longer produce audible speech.

## Code availability

All source code supporting this study will be made publicly available on https://github.com/cronelab/corticom-neural-vad upon acceptance of the manuscript. Moreover, the repository also comes with a bash script which can be used to replicate all steps done in this study, including rendering the figures and running the real-time BCI on streamed signals.

## Data availability

All data supporting this study will be made publicly available on www.osf.io upon acceptance of the manuscript. Neural recordings are prepared in the MATLAB file format version 5, where time-aligned anonymized acoustic speech is stored in the wav file format.

## Author Contributions

M.A. and N.C. wrote the manuscript. M.A., S.J., S.L., Q.R. and D.C. analyzed the data. M.A. S.L. and Q.R. collected the data. M.A., S.J. and G.M. implemented the code for model training and system design. M.A. and S.J. made the visualizations. C.G., K.R., L.C. and N.M. conducted the medical procedure. F.T. handled the regulatory aspects. N.C., N.R. and M.F. supervised the study and the conceptualization. All authors reviewed and revised the manuscript.

## Acknowledgments

Research reported in this publication was supported by the National Institute Of Neurological Disorders And Stroke of the National Institutes of Health under Award Number UH3NS114439 (PI N.E.C., co-PI N.F.R.). The content is solely the responsibility of the authors and does not necessarily represent the official views of the National Institutes of Health.

## Competing Interests

The authors declare that they have no competing interests.

**Supplementary Figure S1.**
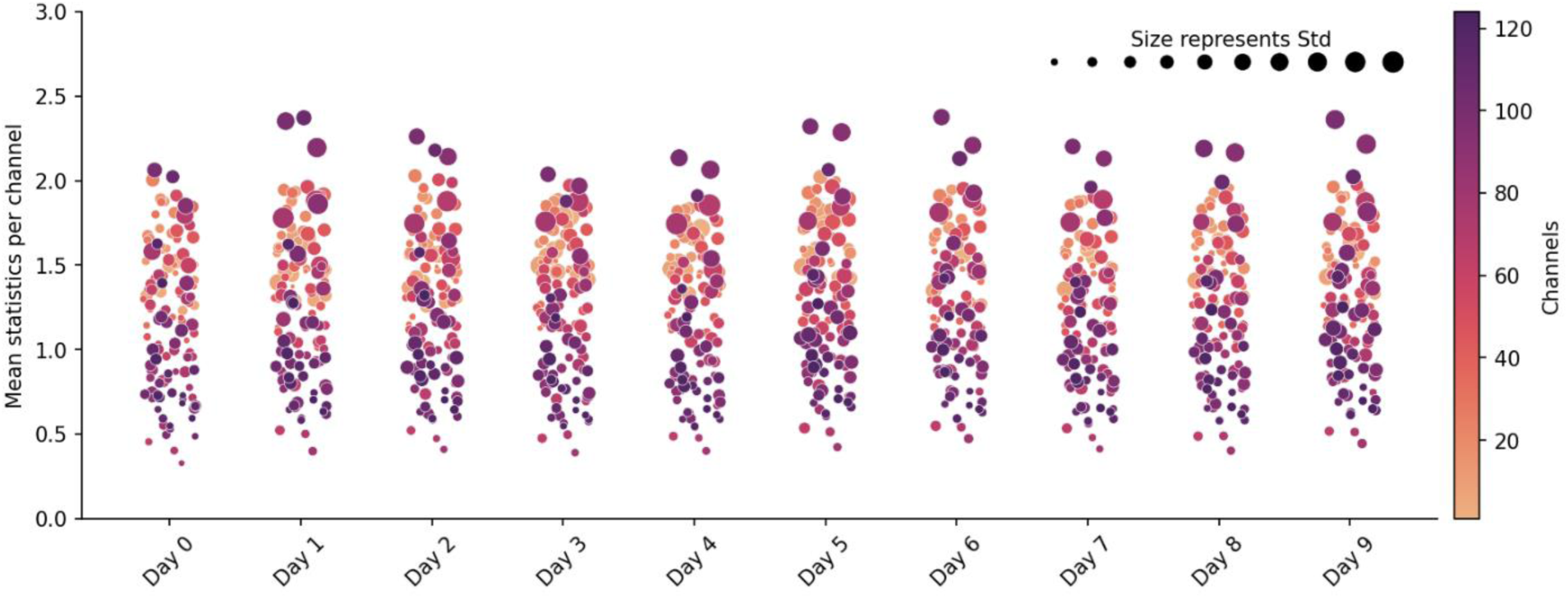
| High-gamma ECoG activity remained stable across all study days. Similar structures can be observed for the majority of the channels, indicating that those channels had comparable activity values and were stable during the study period. Randomized channel shifts on the x-axis were conducted using the same noise profile for all days to only encode relevant information with respect to the y-axis, size and color. Bad channels 19, 38, 48 and 52 were omitted in this plot.

**Supplementary Figure S2.**
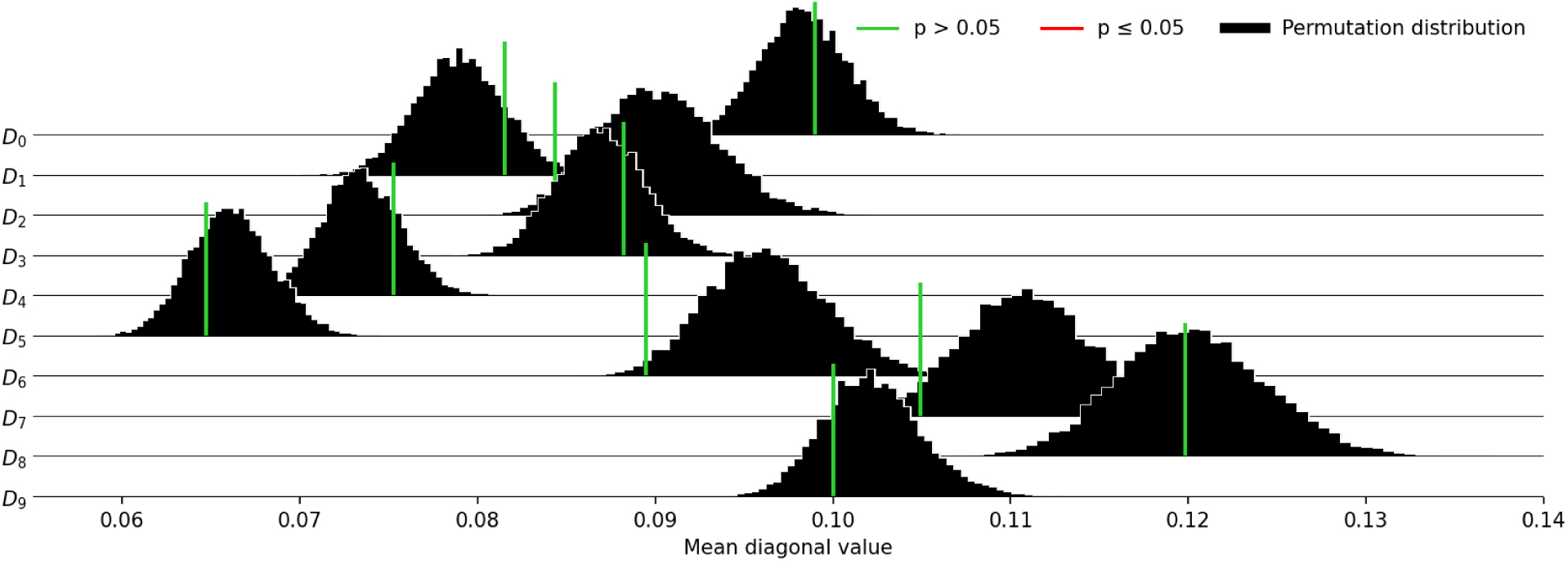
| Summary of the acoustic contamination report. All recordings within our development set (D_0_) and the days for the open-loop recordings (D_1_ – D_9_) were checked for acoustic contamination by using Roussel‘s method^37^. Each histogram visualizes for one day the distribution of mean diagonal values from permutated contamination matrices (N=10,000 permutations). The vertical-colored bars represent the actual mean diagonal value of the contamination index. The statistical criterion for rejecting the null hypothesis is either displayed in green (p > 0.05) or red (p ≤ 0.05) indicating that the neural signals have been acoustically contaminated. We observed in one channel contamination artefacts for one day (day 7) and replaced those high-gamma values with mean activity from neighboring channels. After this step, no acoustic contamination was present anymore. All recordings from the closed-loop block were omitted here as they have not been used for model training.

**Supplementary Figure S3.**
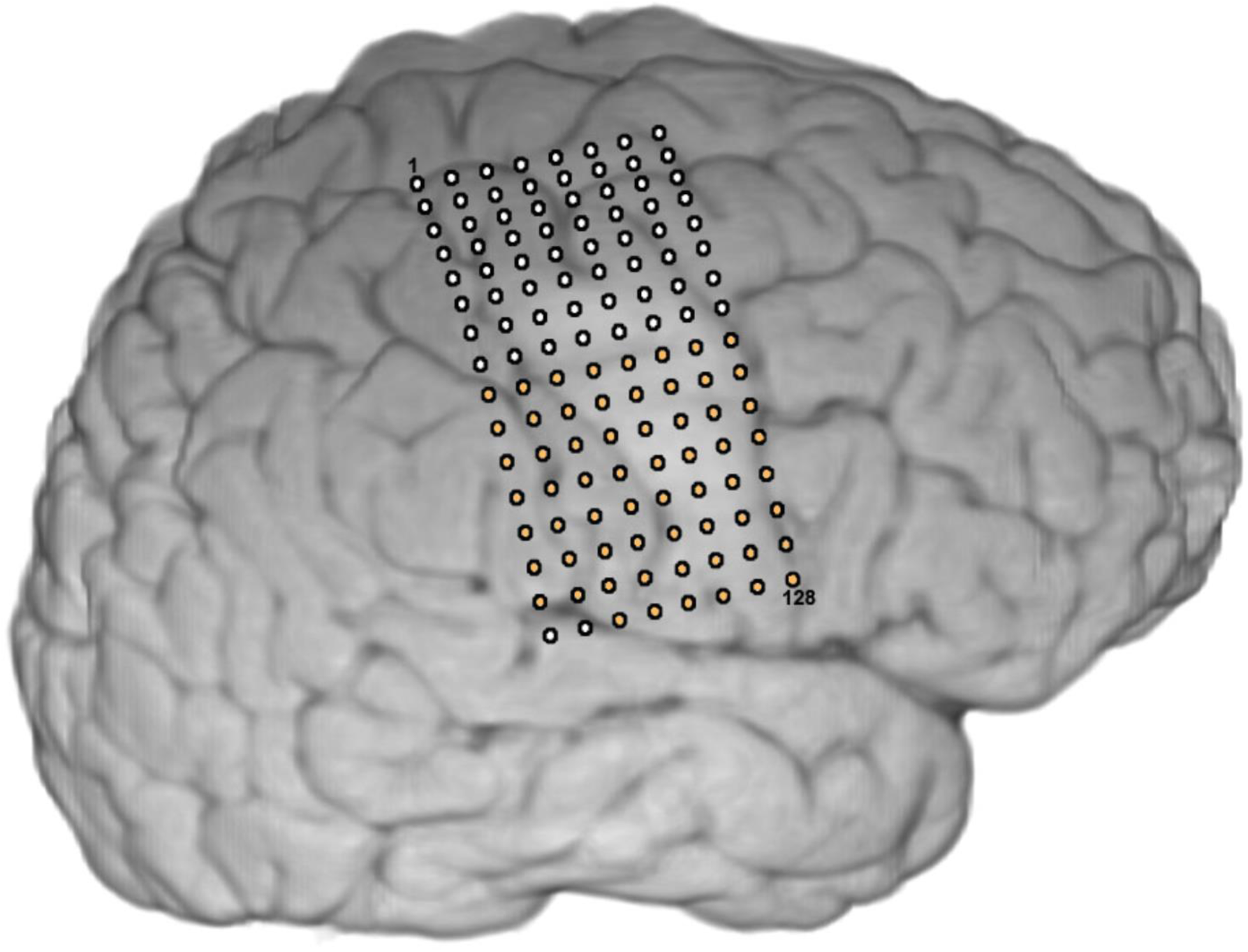
| ECoG array placement in an epilepsy patient to infer a suitable hyperparameter configuration. We determined appropriate hyperparameters for the TICC algorithm by using data recorded from an epilepsy patient implanted with a 128 channel ECoG grid covering similar speech areas than our clinical trial participant. We selected the bottom 62 channels (2 electrodes covering superior temporal gyrus were excluded) to roughly match similar areas than our clinical trial participant. Although the ECoG grid in this patient was implanted on the right hemisphere we observed strong high-gamma activity during speech production, supporting similar observations previously reported in the literature^13^.

## Notes

### Competing Interest Statement

The authors have declared no competing interest.

### Clinical Trial

NCT03567213

### Author Declarations

The Institutional Review Board (IRB) of the Johns Hopkins University gave ethical approval for this work and the Food and Drug Administration (FDA) gave approval under an investigational device exemption (IDE)

